# Ocular light and optical radiation exposure as a modifiable environmental determinant of health: Expert consensus on research gaps and priorities

**DOI:** 10.1101/2025.08.27.25333976

**Authors:** Manuel Spitschan, Anna M. Biller, Kai Broszio, Elaine Fischer, Janice Hegewald, Sylvia Rabstein, Elmar Saathoff, Karin Smolders, Salma M. Thalji, Sarah Weigelt, Daniela Weiskopf, Johannes Zauner

## Abstract

Light exposure over 24 hours is a modifiable environmental influence on human physiology and behaviour with significant implications for health and well-being, yet the field lacks coordinated research infrastructure, standardized methodologies and translational pathways. To address this, we convened a multi-disciplinary consensus workshop and expert consultation process with 13 experts from academia, public health, radiation protection and occupational health institutions. The aim was to identify key research gaps and to define priority areas to guide future work. Through an in-person and hybrid meeting, followed by iterative refinement and feedback, we identified nine critical gaps: (1) lack of standardized measurement tools, (2) inadequate exposure estimation infrastructure, (3) inconsistent descriptors and metrics, (4) absence of outcome standards, (5) limited dose-response evidence beyond the laboratory, (6) insufficient data on intervention effectiveness, (7) poor characterization of vulnerable populations, (8) fragmented data harmonization and (9) limited integration into public health frameworks. To address these gaps, we propose 11 research priority areas spanning measurement, methodology, data infrastructure, ethics and implementation, as well as four capacity building priority areas. This agenda provides a strategic foundation for building an integrated and evidence-based approach to studying and understanding light exposure as a determinant of health.

## Introduction

Light exposure over the 24-hour cycle is a key environmental signal that plays a critical role in regulating human biology, behavior and health. Beyond its well-established function in enabling vision, light influences a range of non-visual physiological processes. These include the synchronization of circadian rhythms and modulation of sleep-wake cycles, hormonal regulation, mood, alertness and metabolic function [1]. Recent advances from in-laboratory research and field assessments have highlighted the importance of understanding how patterns of light exposure (defined by intensity, spectrum, timing, duration and spatial distribution [2]) drive these physiological processes.

In parallel, there is growing evidence that “suboptimal” light exposure and lighting environments, both indoors and outdoors, may contribute to adverse health outcomes. Suboptimal light exposures include insufficient daytime light exposure and limited access to daylight and excessive light exposure before sleep or at night. Emerging efforts to collect continuous light exposure data in diverse populations [3, 4] indicate that both daytime and pre-sleep exposure falls largely outside of current, lab-derived recommendations for healthy light exposure [5]. These recommendations suggest a melanopic equivalent daylight illuminance (melanpic EDI) of >250 lux during daytime, <10 lux in the pre-sleep environment and <1 lux in the sleep environment. For comparison, indoor lighting typically ranges between roughly 10 and 1000 lux melanopic EDI, while outdoor light exposure can reach levels of up to approx. 100,000 lux melanopic EDI [6]. Actual light exposure under real-world conditions [7] values can differ widely, with average values being lower than the possible maximum values [8, 9].

Recent large-scale studies link excessive light exposure at night and dim daytime light exposure to negative health outcomes [10–13]. Preceding these large-scale studies, light at night was assessed by the International Agency for Research on Cancer as a potential factor in the possible development of cancer through night work [14]. These effects may be influenced by urbanization, increased screen use and occupational changes, but they are not well captured in population health data or integrated into routine public health practice. While the scientific evidence supporting the relevance of light exposure is expanding, progress in translating this knowledge into large-scale research, public-health surveillance and policy has been slow. Several challenges contribute to this gap. These include the absence of standardized measurement tools, limited data infrastructure, unclear dose-response relationships and a lack of coordinated efforts to identify, study and protect understudied and vulnerable populations. Furthermore, there are only a few established pathways to translate current research knowledge into practice and incorporate light-related considerations into healthcare systems, occupational safety frameworks, building and lighting design, or public health guidelines.

To address these limitations and to support evidence-based actionable strategies, a structured expert consensus process was initiated. The goal was to identify key research gaps and define priority areas to guide the development of tools, methods and policy relevant to light exposure and health *at scale*. This report presents the outcomes of that process, offering a research agenda for the next decade to advance the field of light and health in a coordinated and impactful way.

## Methods

### Consensus development process

The identification of research gaps and priorities was conducted through a structured expert consultation process combining workshop discussions and iterative written feedback.

### Participants and selection criteria

Thirteen experts (all co-authors of this paper) participated in the development process. Experts were purposively selected to ensure broad coverage of disciplinary domains relevant to non-visual effects of light, including chronobiology, vision science, public health, lighting engineering, radiation protection, and occupational health. Eligibility was based on demonstrated domain expertise (e.g., peer-reviewed publications, contributions to standards or guidelines, or applied professional experience). To facilitate in-depth discussion and iterative consensus building, the group size was intentionally limited to thirteen participants, spanning career stages from graduate student to full professor. All experts were affiliated with European institutions; as such, while the group reflects disciplinary diversity, it does not represent a comprehensive geographic or demographic distribution.

### Hybrid workshop

A one-day hybrid workshop was held on 26 June 2025 at the TUM Institute for Advanced Study (TUM-IAS), Garching, Germany. The agenda included short impulse talks to provide context, followed by breakout group discussions and plenary sessions. Thematic groups identified candidate research gaps and opportunities based on disciplinary expertise and shared observations.

### Synthesis and consultation

Notes and outputs from the workshop were synthesized by the first author into a preliminary draft of research gaps and priorities. This draft was circulated to all participants in two iterative rounds of written consultation. Participants provided detailed comments, corrections and additions, which were incorporated into successive revisions.

### Agreement and consensus

Consensus was defined as the absence of outstanding objections after the final consultation round. Disagreements were resolved through an open commenting process. All participants reviewed and approved the final version. This structured consensus workshop and consultation process generated a shared agenda that reflects the perspectives of a multidisciplinary group of experts, providing a coordinated foundation for future research.

## Results

The expert consultation process identified nine research gaps, as well as eleven research priorities areas and four capacity-building priority areas aimed to fill these gaps.

### Identified research gaps

An overview of the identified research gaps with a description and their relevance for practice can be found in Table 1.

**Table 1:** Gaps in research and practice.

| Gap | Gap description | Why it matters |
| --- | --- | --- |
| **Gap 1**<br>Lack of standardized measurement tools | There are currently no validated, harmonized instruments to monitor and quantify personalized, physiologically- and behaviourally relevant light exposure in the field over long periods of time, including UV/NIR wavelengths and dimensions of visual experience and tools to quantify, assess and evaluate exposure and data quality | Without standardized tools, findings cannot be compared or pooled across studies, limiting scientific progress and evidence-based recommendations |
| **Gap 2**<br>Inadequate exposure estimation infrastructure | There is no robust methodology for estimating light exposure, light availability and other aspects over the 24-hour day using job-exposure matrices, GIS/time-use models or electronic health records | Reliable estimation tools are essential for evaluating typical exposures at the population level where direct measurement is impractical |
| **Gap 3**<br>Lack of consistent exposure descriptors and exposure metrics | There is no consensus agreed-upon list of exposure descriptors or terminology for light-related exposures | This inconsistency fragments the field, prevents harmonization of datasets and makes meta-analyses of published findings very difficult |
| **Gap 4**<br>Absence of core outcome standards | There are no agreed-upon minimum outcomes or protocols to consistently evaluate the health effects of light exposure | This prevents meaningful comparisons across studies, hinders evidence synthesis and weakens policy guidance |
| **Gap 5**<br>Limited dose-response evidence | There is little understanding of how much change in light exposure leads to which health benefit or risk in real-life situations and how much this varies alongside different population and location axes (age, health status, socio-economic status, etc.) | Dose-response data are critical for setting actionable, evidence-based guidelines and recommendations |
| **Gap 6**<br>Limited evidence on intervention effectiveness | There are few studies evaluating light-based interventions placed in the behavioural real-world context for feasibility, cost-benefit and real-world implementation | Without this evidence, policy and practice cannot confidently scale or invest in light-based interventions |
| **Gap 7**<br>Poor characterization of globally representative and vulnerable populations | There are very few studies on light exposure across the globe and research has not adequately identified which subgroups are most vulnerable to light-related health effects, nor their windows of susceptibility | Without this knowledge, interventions and policies may be inequitable or fail to protect those most at risk |
| **Gap 8**<br>Fragmented data harmonization and sharing | No coordinated, FAIR-compliant, federated data-sharing infrastructure exists for light exposure data, including standardized metadata schemas for device properties, protocol-level context, structured exposure descriptors or ontologies and linkages to modeled exposures | This makes collation across studies labor-intensive and error-prone, limits reproducibility and undermines collaboration, automation and large-scale evidence synthesis across different cultures and geographical regions |
| **Gap 9**<br>Limited integration of non-visual effects of light into public health frameworks | Despite growing evidence, the non-visual effects of light are not systematically integrated into public health guidelines or building standards and there is limited awareness in the general public related to the potential impact of light on human functioning, beyond vision | This limits the implementation of research findings in practice, particularly in the areas of urban planning, lighting in schools and workplaces and healthcare. |

### Exposure measurement and estimation

#### Gap 1: Lack of standardized measurement tools

A central limitation in advancing research on light exposure and health is the absence of standardized tools for measuring personal ocular light exposure in real-world settings at scale. Currently available devices differ substantially in terms of spectral sensitivity, dynamic range, angular response and sampling frequency and most importantly, form factor and thus wearability [15–20]. Few of them have been validated against traceable calibration standards and many do not measure across the full photobiologically relevant range of wavelengths, including ultraviolet (UV) [21–23] and near-infrared (NIR) light. Even devices which provide accurate spectral readings according to current standards do not necessarily measure correctly in terms of human physiology, which is especially the case for their measurement field of view and angular response [24–26]. A key limitation of wearables is that wearing location can influence the measured light exposure significantly. Wrist-worn light loggerse can be easily covered by clothing or bedding [27–31]. At the same time, light loggers placed in the corneal plane (e.g., [32–36]) are not readily available for use in every-day contexts [37, 38]. Most wearables and loggers do not capture contextual information related to visual experience, such as the direction of gaze, contrast or scene composition, which may modulate physiological responses [2, 39]. These inconsistencies limit the comparability of data across studies and make it difficult to synthesize findings or draw reliable conclusions from pooled datasets.

In addition to variation in devices, there is a lack of standardized protocols for data collection (e.g., [40]), quality assurance and reporting. Studies often differ in how and where devices are worn, for how long they are used and how missing or noisy data are handled. Very few publications report uncertainty estimates or provide metadata related to calibration and performance characterization [26]. Without transparent and harmonized procedures, reproducibility remains a challenge and collaborative efforts across sites become difficult to implement. The field urgently needs openly available, validated measurement toolkits that include device specifications, standard operating procedures, metadata templates and quality control guidelines that should be adaptable to various use cases. To ensure broad applicability, these resources should be designed to meet the needs of diverse populations including children, older adults, shift workers and clinical patients. Developing and disseminating such tools is the foundation for producing robust, generalizable knowledge about the impact of light exposure on health.

### Gap 2: Inadequate exposure estimation infrastructure

Wearable devices offer one method for capturing individual light exposure. In contrast, large-scale studies and population surveillance efforts often require methods that do not rely on direct measurement, but rather on self-reported or expert-based information on exposures or other data sources Currently, there is no robust infrastructure for estimating personal light exposure or light availability using indirect approaches, such as environment-bound sensor measurements, job-exposure matrices (JEMs), geospatial information systems (GIS), satellite data, time-use models, building automation systems (occupancy records and stationary sensors), or data from electronic health records (EHRs). These methods are well established in other areas of environmental exposure research, such as air pollution and noise [41–44]. As a result, we lack scalable approaches to estimate typical light exposure patterns across diverse geographic regions, occupations and population groups to model population-level or sub-group risks or to identify areas in which interventions could be most beneficial.

To make meaningful progress, the field needs validated methods for indirect exposure estimation that can complement or (partly) substitute wearable data, especially in settings where direct measurement is not feasible due to logistical, financial or ethical constraints. This includes developing light-specific job-exposure matrices that capture occupational lighting conditions, GIS-based models that incorporate building orientation, light-related design specifications, local daylight availability and outdoor behavior and movement patterns and integration of lighting-related variables into EHR or cohort metadata. In the absence of such tools, researchers must rely on assumptions or incomplete proxies that may not reflect real-world conditions. Developing exposure estimation tools that are open, scalable and grounded in empirical data with known accuracy will be key for enabling epidemiological studies and for informing public health surveillance systems that include light as an environmental determinant of health.

### Gap 3: Lack of consistent exposure descriptors and exposure metrics

A barrier to progress in light and health research is the absence of a shared framework for describing and quantifying light exposure. While there is a defined and widely accepted metric on the stimulus strength of light on the non-visual system (CIE S 026/E:2018 [45]), this metric alone only captures momentary intensity and spectral composition and is primarily suitable for laboratory experiments with highly controlled settings. There is currently no consensus on which descriptors or metrics are most relevant for capturing other biologically meaningful aspects of light, such as temporal patterns, spatial distribution, or timing of exposure relative to biological rhythms [19]. Different studies often use different metrics, including mean photopic illuminance (lux), irradiance, melanopic equivalent daylight illuminance or novel metrics that condense time series into singular quantities or aggregated values per specific time windows (like 30-minute or 60-minute bins), mostly without clear justification or standardization [19]. This lack of consistency and guidelines to quantify light in real-life settings fragments the field, reduces the comparability of results and complicates efforts to synthesize findings across published studies and populations.

Moreover, existing descriptors often fail to capture the full complexity of visual and non-visual light exposure in real-world contexts. Metrics may ignore angular geometry, spatial variation across the visual field [25, 46] or the dynamic nature of light exposure due to movement, gaze direction or scene changes throughout the day [2, 39]. There is also a lack of clarity regarding which metrics are appropriate for which research questions, making it difficult to interpret findings or develop guidelines based on exposure thresholds (such as those developed from laboratory studies [5]). Establishing a minimal set of core descriptors, supported by mechanistic evidence, linked to health outcomes and aligned with international standards, will be essential for harmonizing datasets and enabling coordinated progress across research groups, settings and disciplines.

### Health outcomes and mechanistic understanding

#### Gap 4: Absence of core outcome standards

A key challenge in the field of light exposure and health is the lack of agreed-upon core outcome standards. While a growing number of studies have explored how light affects sleep, circadian rhythms, sleepiness, mood, cognition, metabolic function and other health-related endpoints, there is no common set of outcomes or assessment protocols used across studies [47, 48]. Further, the non-visual system affects many downstream physiological health-related functions in a meaningful way, but seldom is the primary determinant, which requires insights into a multitude of research disciplines. Researchers select outcomes based on disciplinary conventions or local study practice and aims, leading to wide variability in what is measured, how it is measured and when measurements are taken. This heterogeneity makes it difficult to compare findings, combine datasets, or conduct robust meta-analyses. It also hampers the translation of evidence into policy and practice, as there are no unified indicators that can serve as benchmarks for intervention studies or public health monitoring.

There is an urgent need to define a core set of outcomes that are mechanistically justified, clinically relevant and feasible to measure in both research and applied settings. Such a standard set would improve reproducibility, support evidence synthesis and create a stronger basis for integrating light exposure considerations into clinical and public health frameworks.

### Gap 5: Limited dose-response evidence

Despite increasing interest in the health effects of light exposure, there is still limited evidence describing the dose-response relationships between light and specific physiological, behavioural and/or clinical outcomes such as metabolic or cardiovascular health [49]. Most existing dose-response studies examine short-term physiological outcomes conducted under tightly controlled laboratory conditions [5, 50–52], which may not reflect real-world exposure patterns or variability in individual responses [53, 54]. As a result, it remains unclear how much light, of what type, at what time of day and for how long, is needed to produce meaningful health effects. Questions about threshold levels, saturation points and the shape of dose-response curves under real-world conditions are largely unanswered [55]. This limits the development of clear, evidence-based guidelines for healthy light exposure across settings such as workplaces, schools, hospitals, care facilities, or urban environments. While there is strong evidence that non-visual effects of light exposure are significant in the laboratory, the size of these effects in the real world is likely to be at best of the same magnitude as the effects seen in the laboratory, at worst washed out by other lifestyle factors and likely lower than those seen in the laboratory [56–60]. Recent large-scale studies relating light exposure to health outcomes suffer from low measurement fidelity due to use of wrist light loggers with a limited range, and require statistical transformations for effective use in predicting health outcomes [10, 12, 13, 61].

Another important gap concerns individual-level variation in dose-response relationships. Individual variability in the response to light is well-established [53, 54, 62]. Factors such as age, sex, gender, health status, chronotype, occupation, geography and socio-economic background may all influence how individuals respond to light exposure, yet these modifiers are rarely accounted for systematically [63]. Additionally, many studies examine single endpoints in isolation, without considering trade-offs or interactions between outcomes such as alertness, sleep, or circadian alignment. There is a need for large-scale, multi-site studies that integrate controlled experimental data with longitudinal observational data in the same individual to map dose-response functions in real-world contexts. Developing predictive models that incorporate individual and environmental variability will be essential for setting practical exposure targets and designing effective and tailored light-based interventions [64].

### Interventions and implementation

#### Gap 6: Limited evidence on intervention effectiveness

Although numerous laboratory studies have shown that ocular light can influence sleep, circadian rhythms, alertness, mood and ocular health, there is limited evidence on the effectiveness of light-based interventions when implemented in real-world settings. Interventions such as circadian lighting systems, daylight-enhancing architectural designs, or behavior change programs are often tested in controlled environments or pilot studies, with limited attention to feasibility, adherence or long-term outcomes. Building-related interventions are most often based on simulations and spot measurements, making their actual effect on personal light exposure ambiguous [64]. This creates a gap between mechanistic understanding and practical application. The few examples, e.g. for behavior change programs targeting the influence of ocular light exposure on eye health are, however, very promising and offer first insights into the feasibility of such long-term interventions (References). Robust implementation studies are essential to understand whether these interventions are effective in real-world settings, for whom and under which conditions. Without them, important contextual challenges and practical barriers may remain unrecognized. Moreover, few studies examine the acceptability, cost-effectiveness, or sustainability of light-based interventions. End-user perspectives are rarely included in the design and evaluation process and potential barriers such as geolocation, cultural preferences, architectural constraints, limited access to daylight and different views on daylight are often overlooked [65]. There is also a limited number [59, 66, 67] of comparative studies evaluating different types of interventions against each other or against existing standards of care. To move forward, the field needs pragmatic trials and real-world implementation research incorporating behavioral, economic and environmental factors. This includes measuring not only physiological, performance-based and health-related endpoints but also usability, equity and scalability across diverse settings such as schools, healthcare facilities, offices and homes.

### Gap 7: Poor characterization of globally representative and vulnerable populations

Many studies on light exposure and health have been conducted in relatively homogenous samples from Western, educated, industrialized, rich and democratic (WEIRD) countries [68], often consisting of healthy young adults in controlled settings. As a result, there is limited understanding of how different population groups may vary in their sensitivity to light-related health effects. For example, a recent study showed different circadian gene expressions in arctic residents, depending on whether they were indigenous or not [69]. Potentially vulnerable populations, such as children, older adults, shift workers, individuals with chronic illnesses, people with disabilities and those living in low-light or high-exposure environments, may respond differently to the same light exposure. A recent study [70] showed that individuals between the ages of 40 and 60 undergo substantial nonlinear changes in their biological profiles during middle and older adulthood. These transitions occur during productive working years, suggesting these periods are critical for intervention. Therefore, recognizing and addressing these changes could be crucial for occupational medicine, as they may affect health outcomes and work ability.

Different target populations may also encounter different types of constraints in accessing daylight or avoiding harmful light at night. However, few studies have systematically identified who is most at risk, when susceptibility is greatest or how vulnerability intersects with environmental, occupational, or social factors.

In addition, there is a lack of data on how individual characteristics such as chronotype, pigmentation, hormonal status (including menstrual cycle or pregnancy) or pre-existing sleep and mood disorders might modulate light sensitivity. Social determinants of health, including income, housing conditions and occupational setting, are rarely considered in light exposure studies, even though they likely influence both exposure and health outcomes. Without better characterization of vulnerable populations and their specific exposure contexts, interventions may be inequitable or ineffective. Future research must prioritize subgroup analyses, stratified study designs and participatory approaches that include the perspectives and needs of underrepresented groups. This will be essential for designing targeted, inclusive and effective public health strategies related to light exposure.

### Infrastructure, data sharing and integration

#### Gap 8: Fragmented data harmonization and sharing

A critical limitation in light exposure research is the lack of coordinated infrastructure for data harmonization, interoperability and sharing. Although a growing number of studies collect wearable light exposure data, the absence of shared metadata standards, ontologies and reporting conventions makes it difficult to compare or combine datasets, a common problem in many fields of science [71, 72]. Device specifications, sampling protocols, placement conventions, spectral sensitivities and pre-processing steps are often reported inconsistently or not at all. This lack of standardization reduces reproducibility and hinders collaboration across institutions, disciplines and countries. As a result, researchers often need to invest substantial time and effort into reprocessing or reformatting data to enable basic comparisons and meta-analyses become labor-intensive and error-prone.

Furthermore, there is no established, FAIR-compliant (Findable, Accessible, Interoperable, Reusable; [73]) platform for storing and sharing light exposure data in a secure, structured and privacy-conscious manner. Many datasets remain inaccessible, “available upon (reasonable) request” [74], or otherwise gated (e.g., login or licensing requirements). There is also limited tooling for merging light exposure data with other health-relevant information, such as actigraphy, environmental conditions or health records. To address these issues, the field urgently needs shared infrastructures, including standardized metadata schemas [75], controlled vocabularies for exposure descriptors, standard operating protocols for data collection and pre-processing, platforms for federated data access, and reproducible analysis pipelines. Enabling interoperable and scalable data sharing will not only improve transparency and reproducibility but also support large-scale evidence synthesis and cross-cultural comparisons in light and health research.

### Gap 9: Limited integration of non-visual effects of light into public health frameworks

Although the scientific understanding of the non-visual effects of light has advanced considerably, this knowledge has yet to be meaningfully integrated into public health frameworks, occupational health guidelines or building and lighting standards. Light is still primarily regarded as a visual requirement, with regulatory and design frameworks focused on visibility, safety and energy efficiency, rather than on circadian, hormonal, affective, or cognitive outcomes. Despite growing evidence that light exposure can influence sleep, mental health, metabolic function and overall wellbeing [10, 11, 13, 76], these effects are not yet routinely considered in urban planning, education, workplace health or healthcare infrastructure. As a result, opportunities to use light as a health-promoting environmental factor remain underutilized.

Public awareness of the health-related impacts of light beyond vision also remains limited. Most individuals are unaware of how daily light exposure patterns affect their sleep, alertness and internal timing and guidance on healthy lighting is rarely included in clinical practice, health education or health promotion materials. In parallel, there are few institutional mechanisms for translating research findings into actionable recommendations or structural interventions, such as lighting policies in schools, hospitals, or elder care facilities. Bridging this gap will require close collaboration between researchers, policymakers, designers, end users and public health professionals. Clear, evidence-based communication, alongside the development of practical tools and training materials, will be essential for embedding non-visual light considerations into public health practice at scale.

### Identified priority areas and recommendations

The expert group identified a series of priorities for research and practice designed to address the gaps outlined above, these can be found in Table 2.

**Table 2:** Priority research areas.

|  | Gaps | Priority area | Short-term (0-18 months) | Medium-term (18m-5y) | Long-term (5-10y) |
| --- | --- | --- | --- | --- | --- |
| **Exposure** | 1, 2,3 | Development and refinement of exposure measurement tools | Develop and validate brief questionnaires in multiple languages; visual experience assessments and UV/NIR | Pilot these tools in diverse populations and refine based on feedback | Integrate validated gold-standard toolkits into national and international health studies |
|  |  | Development of exposure estimation methods | Develop job-exposure matrices and initial GIS/time-use models; stationary measurements | Validate and benchmark estimation models with wearable data | Integrate estimation tools into EHR systems and surveillance frameworks |
|  |  | Definition of exposure descriptors and exposure metrics | Convene expert panels to agree on a minimum list of exposure descriptors in an evidence-based way | Coordinate with international partners (e.g., CIE, ISO) to align descriptor terminology | Promote these descriptors as international reference standards for data collection |
| **Outcomes** | 4, 5 | Definition of core outcome sets | Convene expert panels to define protocols and minimum outcomes with known mechanistic evidence | Validate (standard) methods/protocols through multicenter studies | Institutionalize protocols in journals, funding calls and public health guidance |
|  |  | Definition of dose-response relationships | Map existing evidence on light dose-response relationship | Conduct controlled experiments and cohort studies to quantify dose-response relationships; develop predictive models | Integrate validated dose-response functions into national/international health guidelines |
| **Interventions** | 6 | Definition of interventional targets | Identify promising light-based interventions (e.g., circadian lighting, outdoor time promotion, screen hygiene) | Conduct pilot intervention studies; assess implementation feasibility, acceptability, user awareness, health-economic aspects and barriers in real-world conditions | Scale up successful interventions in diverse settings, integrate intervention guidelines into public and occupational health, education and urban design policy |
| **Population** | 7 | Definition of sensitive populations and windows of vulnerability | Identify priority groups and hold co-creation workshops to identify needs and barriers for specific groups | Launch targeted studies with tailored exposure measurement and interventions | Embed subgroup-sensitive standards in routine public health practice; develop and maintain equitable access to research and interventions |
| **Data and analytics** | 1, 8 | Interoperable data and reproducible analyses | Publish reproducible pipelines and initial metadata templates | Train researchers in dataset documentation and harmonization; develop tools to align legacy datasets with shared vocabularies and ontologies. | Maintain and evolve interoperable workflow standards |
|  |  | Data sharing and reference data sets | Launch FAIR-compliant data repository ("Global Light Commons"); define minimum dataset criteria | Populate with data; provide tools and training for curation, licensing and metadata annotation | Maintain a global federated network of interoperable light exposure data hubs, with persistent identifiers, support for cross-repository querying, metadata discovery and dataset versioning. |
|  |  | Ethics and privacy | Map ethical, privacy and equity challenges in light exposure data collection and intervention | Develop and disseminate best practices for secure data handling, structured consent metadata, anonymization and equity-aware data collection | Institutionalize ethics and equity principles in international light exposure research and policy frameworks, through governance checklists, requirements in funding, ethics reviews |
| **Embedding** | 2, 7, 9 | Embedding in public and occupational health | Add light questions to existing national surveys; pilot EHR linkages | Integrate wearable light measurements into ongoing studies; design and a novel dedicated light exposure cohort | Institutionalize light metrics in routine public health monitoring and sustain the dedicated cohort as a reference |

### Scientific and health priorities

#### Development and refinement of exposure measurement tools

To address the lack of standardized light measurement tools, one of the highest priorities is the development and refinement of instrumentation and protocols that can reliably capture physiologically and behaviourally relevant light exposure in daily life and at scale. In the short term, this includes the development and validation of brief self-report tools, such as light exposure and visual experience questionnaires (e.g., [77, 78]), that are easy to administer, adaptable across languages and suitable for population-based surveys. Alongside these, efforts are needed to define minimal requirements for wearable light sensors [15], including spectral range, dynamic range, temporal resolution and calibration standards, particularly for capturing physiologically relevant portions of the spectrum such as melanopic, ultraviolet and near-infrared exposure.

In the medium term, these tools should be piloted in diverse populations and refined based on usability, acceptability and technical performance. Wearables, “passive” logging devices such as smartphones and stationary measurement devices must be evaluated not only for accuracy but also for practicality in free-living conditions, particularly in populations such as young children, older adults and shift workers. Integration of visual experience dimensions (such as spatial contrast, field of view and gaze direction) into measurement approaches will be increasingly important for capturing the complexity of real-world exposures as well as contextual details regarding sensor wear (e.g., registering sensor nonwear), location (indoors/outdoors) and sleep/wake status.

In the long term, the goal is to integrate validated, standardized measurement toolkits into national and international cohort studies, health monitoring systems and environmental surveillance initiatives. These toolkits should be open-source, versionable, modular and interoperable with other data platforms. Establishing reference methods and benchmarking datasets will be essential to ensure comparability across studies, support quality control and accelerate cumulative scientific progress. Investments in this area will lay the groundwork for reliable, large-scale data on light exposure and its effects on human health.

### Development of exposure estimation methods

In many research and surveillance contexts, it is not feasible to collect direct, sensor-based light exposure data from all individuals. Therefore, a key priority is the development of robust methods for estimating individual and group-level light exposure indirectly. In the short term, this includes building light-specific job-exposure matrices (or analogues for different educational settings) that account for typical occupational lighting environments across sectors and roles, as well as initiating geospatial models that integrate factors such as building design specifications, geographic location and daylight availability. Time-use data, where available, could provide additional contextual information on indoor versus outdoor activity, proximity to windows, sleep/wake and typical screen use patterns. These estimation methods should be benchmarked and validated against wearable light sensor data to assess their accuracy and reliability, as well as the target performance required for a specific outcome.

Medium-term efforts should focus on improving model granularity and integrating individual-level modifiers such as age, activity and chronotype. This also includes harmonizing data inputs from sources such as weather databases, satellite imagery, urban planning data and architectural designs. Validation efforts should prioritize both general population samples and groups with distinctive exposure patterns, such as shift workers, schoolchildren, or people in institutional settings.

In the longer term, light exposure estimation models should be embedded in broader environmental health surveillance and linked to electronic health records and cohort datasets. This will enable large-scale exposure-outcome analyses and allow for the estimation of cumulative exposure profiles over time. As in other exposure assessments [79], tools for automated, location-based exposure mapping (e.g., [80, 81]) will be essential for understanding spatial and temporal patterns of light exposure across populations and informing urban and occupational health policy.

### Definition of exposure descriptors and exposure metrics

Establishing a common language for describing light exposure is essential for harmonizing research across studies, disciplines and countries. In the short term, expert panels should be convened to identify a core set of exposure descriptors that are both biologically meaningful and practical to implement. These descriptors should include spectral, temporal, spatial and contextual dimensions of exposure, such as intensity, duration, spectral composition (e.g., melanopic, photopic), timing relative to sleep and wake cycles and angular distribution within the visual field. The selection of metrics should be guided by mechanistic evidence and data and linked, where possible, to known physiological pathways or health outcomes.

Medium-term priorities include aligning terminology and definitions with international standards and regulatory bodies, such as the International Commission on Illumination (CIE), the International Organization for Standardization (ISO), the International Commission on Non-Ionizing Radiation Protection (ICNIRP), the International Labour Organization (ILO) and relevant health authorities. Consistency across fields, ranging from lighting design and chronobiology to epidemiology, public health and health psychology, will facilitate interdisciplinary collaboration and ensure that metrics are broadly understood and applicable. Descriptors should also account for contextual factors, including gaze direction, indoor versus outdoor location and proximity to surfaces or screens. In the long term, these descriptors should be promoted as reference standards for data collection, documentation and reporting. Adoption in cohort studies, clinical protocols, wearable device software and metadata templates will enhance comparability and enable cumulative science. Establishing and disseminating practical guidance documents, including worked examples, case studies and open-source tools for metric calculation, will support consistent implementation and widespread uptake across the research community.

### Definition of core outcome sets

To improve comparability and synthesis across studies, there is a pressing need to define core outcome sets [47, 48] for research on light exposure and health. In the short term, expert working groups should be convened to develop consensus on a minimum set of outcomes that are relevant, mechanistically justified and feasible to assess in both research and applied settings. These core outcomes should span physiological, behavioral and clinical domains and include metrics such as sleep timing and quality, subjective alertness and cognition, mood, metabolic health and circadian (phase) markers. Outcomes should also be adaptable for use in different populations and settings, including children, older adults and individuals with chronic conditions.

Medium-term priorities include validating the proposed outcome measures and associated protocols through multicenter and cross-cultural studies. This should involve assessing the reliability, sensitivity and specificity of outcome tools, as well as their responsiveness to changes in light exposure. Harmonization with outcomes used in sleep medicine, chronobiology, metabolic and environmental health research will facilitate integration into existing evidence bases. Consideration should also be given to the burden of measurement on participants and the practicality of implementation in large-scale or resource-limited studies.

In the long term, the goal is to institutionalize these core outcome sets through their adoption in clinical trial registries, funding calls, cohort designs and journal reporting guidelines. Establishing shared protocols will facilitate evidence synthesis, reduce research waste and support more consistent translation of findings into clinical and public health practice. Widely endorsed outcome standards will also help build the case for including light exposure in routine health assessments and population monitoring systems.

### Definition of dose-response relationships

Understanding how changes in light exposure translate into changes in health outcomes is essential for developing evidence-based guidelines and interventions. In the short term, efforts should focus on systematically mapping existing evidence on dose-response relationships between light and key physiological, affective, cognitive, or clinical endpoints. This includes reviewing experimental and observational studies to identify established thresholds, saturation points and effect sizes across different light parameters, such as intensity, spectral and spatial composition, timing and duration. Identifying gaps in the current literature will help prioritize new studies and refine hypotheses.

In the medium term, dedicated experimental and longitudinal studies are needed to quantify dose-response functions in real-world contexts. These studies should be designed to assess both acute and more long-term effects of light exposure and should include diverse participant groups to account for interindividual variability related to for instance, age, sex, gender, chronotype, health status and socio-economic background. Predictive models that incorporate biological timing, temporal dynamics and contextual variables should be developed to better understand how light interacts with human physiology over time, in real-life settings.

The long-term objective is to integrate validated dose-response functions into public health and clinical guidelines. This will require strong evidence linking specific exposure patterns to measurable health benefits or risks, as well as tools to translate scientific findings into actionable thresholds for different settings (such as homes, workplaces, schools and healthcare environments). Standardizing dose-response models and embedding them in digital tools and health surveillance systems will support risk assessment, behavioral recommendations and intervention planning across populations.

### Definition of interventional targets

To translate knowledge of light exposure into meaningful health benefits, there is a need to identify and define promising interventional targets. In the short term, this involves synthesizing existing evidence to pinpoint light-based interventions that are both biologically plausible and practically implementable. Examples include the timing and spectral tuning of indoor lighting systems, promoting access to daylight through architectural design, encouraging time spent outdoors while considering geolocation and developing behavioral strategies around both daytime and evening light exposure, or proxies thereof, such as encouraging time spent outdoors. These targets should be framed in terms of both potential benefits and feasibility in everyday settings such as homes, schools, workplaces and healthcare institutions.

Medium-term efforts should focus on pilot studies to assess the effectiveness of these interventions under real-world conditions. This includes evaluating user acceptability, technical feasibility, equity of access and cost-effectiveness across diverse environments and population groups. Implementation research will be critical for identifying barriers to adoption, such as competing priorities in architectural or clinical workflows, as well as potential unintended consequences. Moreover, it is essential to determine both the extent and approach of personalization. Co-design approaches that involve end users in intervention development will help ensure that strategies are culturally appropriate and context-sensitive and fit the users’ needs, capacities and constraints.

In the long term, effective interventions should be scaled up and integrated into public and occupational health strategies, educational campaigns and built environment policies. Evidence-based guidance for intervention design and deployment should be developed and disseminated, along with training materials and practical toolkits for professionals in healthcare, education, lighting design and urban planning. Embedding light-based interventions into broader health promotion frameworks will support systemic change and improve population-level outcomes related to sleep, circadian and metabolic health and overall wellbeing.

### Definition of sensitive populations and windows of vulnerability

Identifying who is most at risk from suboptimal light exposure and when, is essential for designing equitable and effective interventions. In the short term, research should focus on mapping population groups that may have increased sensitivity to light-related health effects, including children, older adults, shift workers, people living in low-light environments, individuals with neurodevelopmental or psychiatric conditions and those with visual impairments. Co-creation workshops involving members of these groups, caregivers, clinicians and community stakeholders can help uncover context-specific needs, barriers and preferences related to light exposure and lighting environments.

In the medium term, targeted studies should be launched to characterize exposure-response relationships, vulnerabilities and modifiable factors in these populations. This includes using wearable sensors, tailored questionnaires and physiological assessments to understand how biological timing, light sensitivity and behavioral routines vary across life stages and health contexts. Studies should also explore the intersection of vulnerability with social and environmental determinants, including housing quality, occupational demands and geographic location. Particular attention should be paid to developmental windows, such as infancy, adolescence and older age, when exposure patterns may have especially strong or lasting effects.

The long-term goal is to embed population-sensitive standards into public health practice and policy. This involves ensuring that guidelines, interventions and surveillance systems account for variability in sensitivity and access to healthy lighting conditions. Tools for equitable exposure monitoring and intervention delivery should be developed and scaled, along with strategies to ensure long-term inclusivity in research and implementation. This approach will help prevent widening health disparities and ensure that vulnerable and underrepresented populations benefit from advances in light and health research.

### Interoperable data and reproducible analyses

The ability to reuse and combine data across studies is foundational for scientific progress, yet the field of light exposure and health research currently lacks the necessary infrastructure to ensure interoperability and reproducibility. In the short term, a key priority is the development and dissemination of reproducible analysis pipelines [82] and initial metadata templates [75]. These should define how data are processed, structured and annotated, including device specifications, placement information, calibration procedures and contextual variables such as activity or location. Sharing these pipelines openly, alongside example datasets, will promote transparency and facilitate capacity-building across research teams.

Standardized statistical analysis processes are crucial for these pipelines, as with data preprocessing for data quality and consistency. Robust methods are key for data cleaning, normalisation and statistical modelling. Transparency and reusability are key to ensure the integrity and reliability of findings in light exposure and health research. Medium-term efforts should focus on training researchers in data harmonization and documentation practices, particularly for wearable light exposure data collected in diverse formats and settings. Tools are needed to align legacy datasets with shared vocabularies, ontologies and standard data models. This will allow older datasets to be brought into alignment with newer efforts and enable long-term integration across studies and disciplines. Collaboration with experts in data science, environmental health and biomedical and health informatics will be essential to develop robust, scalable workflows.

In the long term, interoperable workflows should become a standard component of light and health research. This includes maintaining and evolving a set of community-endorsed data models, ontologies and quality assurance tools, along with clear guidance on data provenance, versioning and reusability. Infrastructure should support the full research cycle, from idea generation, data collection to analysis, sharing and reanalysis. These efforts will not only improve scientific rigor but also enhance the potential for collaborative, multicenter research on light exposure and its impact on health across populations and settings.

### Data sharing and reference data sets

To accelerate progress in light exposure and health research, the field urgently needs coordinated efforts to support data sharing and the development of high-quality reference datasets. In the short term, this includes launching a FAIR-compliant (Findable, Accessible, Interoperable, Reusable; [73]) data repository, tentatively referred to as the "Global Light Commons", that enables secure and structured sharing of light exposure data across research groups and institutions. Minimum dataset criteria should be defined, including requirements for metadata, data quality, device specifications, exposure metrics and contextual variables such as population characteristics, time of year and measurement setting.

In the medium term, efforts should focus on populating the repository with high-quality datasets and creating tools and training materials to support data submission, licensing and curation. Metadata annotation should follow agreed standards and use controlled vocabularies to support cross-study querying and interpretation. Attention should also be paid to ethical and privacy concerns, ensuring that data can be shared in a way that respects participant confidentiality and local data governance requirements.

The long-term vision is to maintain a global, federated network of interoperable light exposure data hubs. These should support persistent identifiers for datasets, cross-repository search capabilities and robust systems for versioning, data provenance and quality control. Reference datasets covering a range of populations, geographical regions and exposure contexts will provide benchmarks for method validation, model development and cross-cultural comparison. A coordinated approach to data sharing will enable cumulative science, support large-scale meta-analyses and facilitate evidence-based recommendations for light-related health policies.

### Embedding in public and occupational health

Integrating light exposure as a factor in public and occupational health requires a deliberate, stepwise approach. In the short term, light-related variables should be added to existing national health and lifestyle surveys. This could include self-reported measures of time spent in natural and artificial light, sleep-wake timing and screen use patterns. Pilot linkages between wearable light exposure data and electronic health records (EHRs) should also be explored to assess feasibility, privacy-related concerns and added value for health monitoring and risk prediction.

Medium-term priorities involve embedding wearable light measurements into ongoing population-based studies and designing a dedicated cohort study focused specifically on light exposure and health. This cohort should be diverse in terms of geography, socioeconomic background and age and should include longitudinal tracking of both light exposure and health outcomes. In occupational health, partnerships with employers and workplace safety agencies can support the inclusion of light-related assessments in routine evaluations and risk assessments, particularly for shift workers and employees in low- or high-light environments.

In the long term, light exposure metrics should be institutionalized as part of routine public health monitoring systems and occupational health surveillance. Standard indicators (such as light-at-night exposure, daytime light sufficiency or light misalignment) should be tracked at the population level alongside other environmental determinants of health. The dedicated light exposure cohort should be sustained as a reference resource for policy development, risk modeling and evaluation of intervention strategies. Embedding light into public and occupational health frameworks will ensure that it is treated not only as a visual consideration, but as a modifiable determinant of health, equity and wellbeing.

### Cross-cutting capacity-building priorities

The cross-cutting capacity-building priorities are presented in Table 3.

**Table 3:** Cross-cutting capacity-building priorities.

|  | Priority area | Short-term (0-18 months) | Medium-term (18m-5y) | Long-term (5-10y) |
| --- | --- | --- | --- | --- |
| **Capacity building** | Global research networks and capacity | Establish a novel global research network | Host annual conferences, working groups, researcher exchanges, develop multi-language materials | Sustain long-term global network coordination |
|  | Cross-sector and interdisciplinary collaboration | Set up funded cross-disciplinary consortia | Maintain translational working groups across sectors | Institutionalize cross-sector alliances and standards collaborations; develop formal policy engagement strategies |
|  | Communication and dissemination | Develop public education messages on light and health; design toolkits for clinicians, teachers and workplaces | Launch public campaigns on healthy light exposure | Institutionalize public-facing education programs as part of health promotion; evaluate effectiveness |
|  | Policy engagement | Map current policy gaps and opportunities for integrating light exposure considerations | Create policy briefs and evidence summaries tailored to policymakers | Embed light exposure metrics and interventions into routine public health policy frameworks; knowledge exchange |

### Global research networks and capacity

Establishing and sustaining dedicated research networks is essential for advancing the field of light exposure and health. In the short term, this means forming interdisciplinary consortia that bring together experts from chronobiology, environmental health, lighting design, data science, epidemiology, health psychology, occupational safety and health, and public policy. These networks should support early-career researchers across disciplines, promote knowledge exchange and facilitate collaborative grant applications and shared infrastructure development. It is important to recognize the need for such efforts to be global in scope, as light exposure and health are influenced by cultural, geographic and socioeconomic factors that vary widely across regions. For example, myopia research has generated extensive datasets in Asian countries, whereas comparable data from European populations remain relatively scarce. Positioning this work within a European perspective could therefore help address critical knowledge gaps and also provide a strong foundation for future European grant initiatives in this area. At the same time, the field must move toward greater global inclusion by actively engaging underrepresented regions such as Asia, Latin America, and Africa, thereby ensuring that emerging frameworks and datasets capture diverse cultural and environmental realities. Several initiatives point in this direction, and should be supported in the future (e.g. global collaborative data collection efforts following a harmonized protocol [40] yielding data sets from understudied locations [3, 83]).

### Cross-sector and interdisciplinary collaboration

Light exposure research intersects with multiple sectors (including healthcare (e.g., psychology, etc.), architecture, engineering (e.g., lighting technology), education, urban planning and occupational safety) and effective solutions require input from all of them. Mechanisms should be established to support long-term collaboration across sectors, such as translational working groups, shared standards initiatives and co-designed intervention trials. These partnerships will ensure that research findings are actionable and aligned with real-world needs.

### Communication and dissemination

Public understanding of how light affects health remains low, and few tools exist to translate complex findings into accessible guidance (e.g., see consensus statements [84]). Communication strategies should include public education campaigns, visual materials for clinicians and educators and targeted toolkits for specific settings such as schools or elder care facilities. Engaging professional organizations and media partners can amplify impact and help normalize light as an environmental health factor.

### Policy engagement

Bridging the gap between science and policy requires clear, evidence-informed messaging and sustained engagement with decision-makers. Priority actions include mapping current policy gaps, developing briefs and summaries tailored to policymakers and participating in standard-setting processes for building design, work environments and public health surveillance. Policy engagement should be proactive and ongoing to ensure that aspects of healthy light exposure are integrated into future health and environmental frameworks.

## Discussion

This research agenda identifies a coordinated set of gaps and priorities that are essential for advancing research on light exposure and health. Light is a modifiable environmental factor with widespread implications for circadian biology, sleep, mood, vision, productivity and long-term health outcomes. As this process has shown, the field still lacks many of the fundamental tools, infrastructures and standards necessary to support robust, interdisciplinary and population-level investigations of light exposure.

One key finding of this agenda is the urgent need for methodological standardization. Without common metrics, descriptors or outcome measures, it remains difficult to compare findings across studies, synthesize evidence, or formulate evidence-based recommendations. This fragmentation also hampers efforts to integrate light into broader public and occupational health frameworks. Priority areas such as the development of interoperable data infrastructure, consensus-based exposure metrics and ethical data-sharing practices will be critical enablers of cumulative science.

Several cross-cutting themes emerged throughout the agenda-setting process. First, the field requires stronger interdisciplinary and cross-sectoral collaboration, particularly between researchers, public health officials, occupational health practitioners, architects, lighting engineers and technology developers. Second, greater efforts are needed to bridge the gap between basic science and implementation, ensuring that findings are translated into practical guidance, regulations and behavior change strategies.

The development of this agenda was itself a model of collaborative, multi-stakeholder engagement. Experts from academic, governmental and applied domains contributed to the identification and prioritization of gaps, helping to ensure the agenda reflects both scientific and real-world needs. While the outputs are necessarily high-level, they are designed to serve as a shared foundation for future project development, funding strategies, infrastructure investments and policy engagement.

Moving forward, the implementation of this agenda will depend on sustained coordination, strategic investment and capacity building. The establishment of an international alliance or task force could help maintain momentum, support working groups on specific priority areas and align efforts across countries and sectors. With these coordinated actions, the field will be well-positioned to realize the full potential of light as a determinant of health – and to do so in a way that is equitable, scalable and evidence-based.

### Limitations

Although the consensus process provided a structured approach for integrating expertise across relevant disciplines, several limitations must be acknowledged in evaluating the scope and generalisability of the outcomes.

The expert group was composed entirely of individuals affiliated with European institutions, which limits the geographic, socioeconomic, and cultural representativeness of the perspectives captured. The study did not include experts from low- or middle-income countries, nor did it aim to reflect demographic diversity with respect to ancestry, race, or mixed heritage. As the selection process focused on disciplinary expertise rather than population representation, the findings reflect the views of a specialised but regionally concentrated group. Future work should broaden participation to include experts and stakeholders from additional world regions and diverse backgrounds to enhance global relevance and applicability.

This work is based on expert consensus rather than direct empirical validation. While consensus methods are valuable for synthesising multidisciplinary knowledge and identifying areas of agreement, the conclusions necessarily reflect expert judgement. In domains where empirical evidence is incomplete, heterogeneous, or rapidly evolving, such judgement-based recommendations should be interpreted as provisional and subject to refinement as new data become available.

The scientific evidence on the non-visual effects of light is advancing rapidly, especially in areas such as wearable sensing, exposure characterization, and longitudinal circadian health research. As a result, the recommendations derived from this consensus reflect the state of knowledge at a specific point in time. Emerging findings may necessitate updates or revisions to ensure that the guidance remains aligned with the most current evidence.

Finally, the consensus process involved only scientific and technical experts and did not include end-users, patient representatives, clinicians, policymakers, or industry stakeholders. Their absence limits the ability to capture practical perspectives related to usability, feasibility, equity, implementation challenges, and real-world applicability. Incorporating a broader set of stakeholders in future work will be essential for ensuring that recommendations are both actionable and responsive to the needs of diverse user groups.

## Conclusion

This agenda presents a coordinated roadmap for advancing research on light exposure and health. Through a structured consensus workshop and expert consultation process, we identified nine key gaps: (1) the lack of standardized measurement tools, (2) inadequate exposure estimation infrastructure, (3) inconsistent exposure descriptors and metrics, (4) absence of core outcome standards, (5) limited dose-response evidence, (6) insufficient data on intervention effectiveness, (7) poor characterization of globally representative and vulnerable populations, (8) fragmented data harmonization and sharing and (9) limited integration of non-visual light effects into public health frameworks. To address these, we propose eleven research priority areas: developing and refining measurement tools and estimation methods; defining consistent exposure descriptors, core outcome sets, dose-response relationships, interventional targets and vulnerable populations; promoting interoperable and reproducible data practices; creating infrastructure for data sharing and reference datasets; addressing ethical and privacy considerations; embedding light exposure into public and occupational health systems. Additionally, four capacity-building priorities were identified: global research networks, interdisciplinary collaboration, strategic communication and policy engagement. Taken together, these actions provide a foundation for cumulative, equitable and translational research that positions light as a core determinant of health.

**Figure 1:**
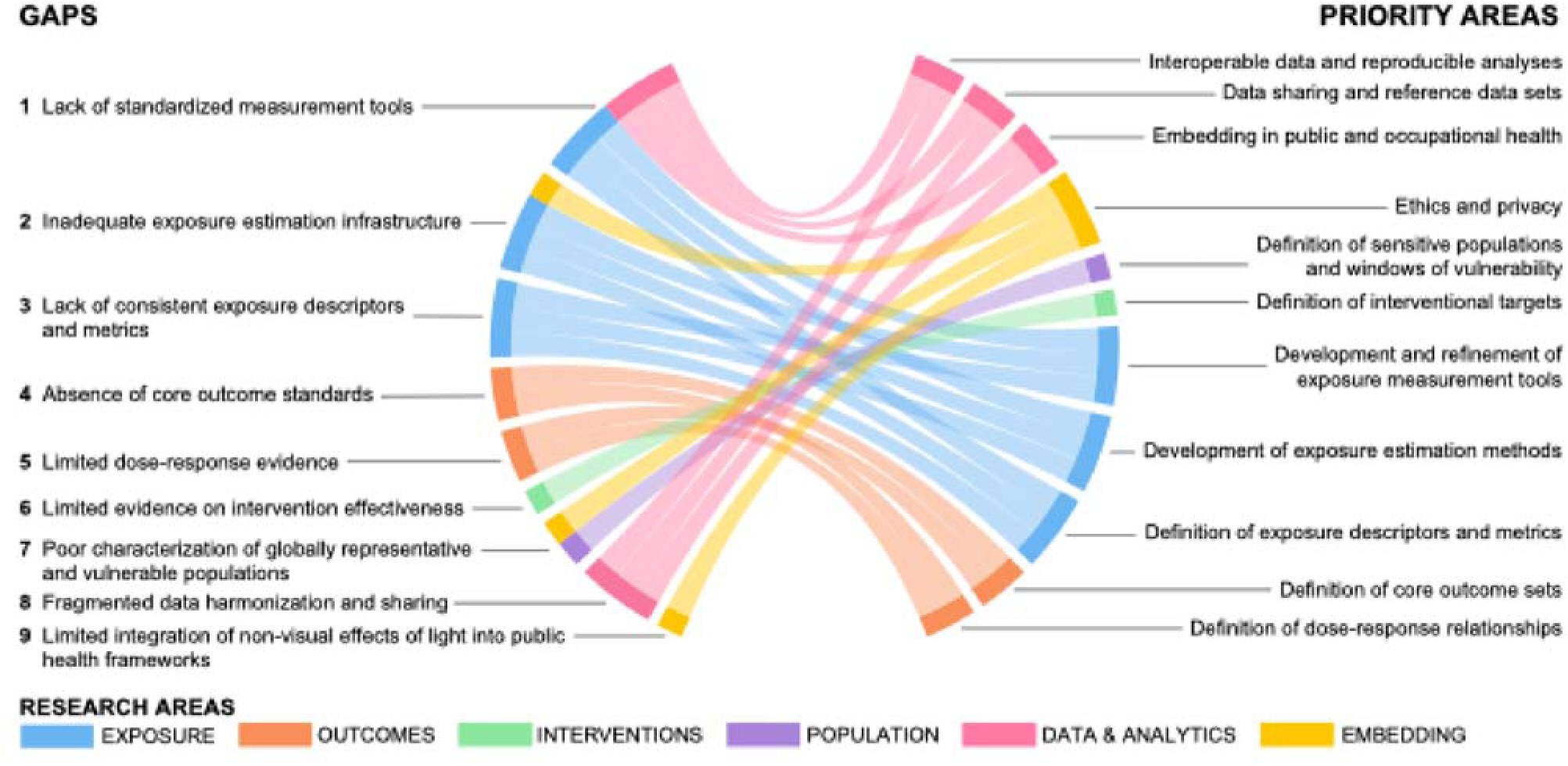
Gaps in research and practice and priority areas.

## Supporting information

Response to reviewers

## Data Availability

No data were generated in this report.

## Statements

### Data availability statement

No data were generated in this report.

### Code availability statement

No code was used in this report.

## Author contributions (CRediT taxonomy)

Conceptualization: MS

Data curation: EF

Formal analysis: –

Funding acquisition: –

Investigation: MS, AMB, EF, ES, DW, JH, JZ, KB, KS, SMT, SR, SW

Methodology: MS, AMB, EF, ES, DW, JH, JZ, KB, KS, SMT, SR, SW

Project administration: MS

Resources: –

Software: –

Supervision: –

Validation: –

Visualization: –

Writing – original draft: MS

Writing – review & editing: MS, AMB, EF, ES, DW, JH, JZ, KB, KS, SMT, SR, SW

Acknowledgments

## Patient and public involvement

Patients and the public were not involved in the design, conduct, reporting, or dissemination plans of this research

## Declaration of generative AI and AI-assisted technologies

During the preparation of this report, the authors used ChatGPT 4o to improve the readability and language of the manuscript. The authors reviewed and edited the content as needed and take full responsibility for the content of the publication.

