## Supplementary material for "Ocular light and optical radiation exposure as a modifiable environmental determinant of health: Expert consensus on research gaps and priorities": Response to reviewers

**Spitschan et al., BMC Medicine**
Response to reviewers

**Editor comments**
We thank the editor and reviewers for their constructive comments on our manuscript. We have revised the manuscript following the comments, and list the changes below.

| **Reviewer 1** | | | |
|---|---|---|---|
| **Comment** | **Reviewer comment** | **Response** | **Related comments** |
| R1.1 | The present paper a strategic foundation for building an integrated and evidence-based approach to studying and understanding light exposure as a determinant of health. I have major comments to this paper. | Many thanks for your comments – we appreciate your thorough reading. | |
| R1.2 | I would like to see a deeper discussion on the difference between outdoor and indoor light exposure. I think the amount of light that enters the eye is important, but recent research seems to be biased towards outdoor light exposure.<br><br>Please cite more of indoor research and deepen your discussion. | We thank the reviewer for this valuable insight. We have expanded this point in the discussion on this:<br><br>*These recommendations suggest a melanopic equivalent daylight illuminance (melanpic EDI) of >250 lux during daytime, <10 lux in the pre-sleep environment and <1 lux in the sleep environment. For comparison, indoor lighting typically ranges between roughly 10 and 1000 lux melanopic EDI, while outdoor light exposure can reach levels of up to approx. 100,000 lux melanopic EDI [6]. Actual light exposure under real-world conditions [7] values can differ widely, with average values being lower than the possible maximum values [8, 9].*<br><br>In the Discussion, we now also include a detailed Limitations section. | |
| R1.4 | I would like to see a more in-depth discussion of the issues with wearable devices. Many large-scale studies in recent years have been conducted using wearable devices, but there has not been sufficient discussion about the issue of the sensor being hidden by sleeves or a blanket. | Indeed, this is an issue of many wearable devices that we have now addressed in the discussion:<br><br>*A key limitation of wearables is that wearing location can influence the measured light exposure significantly. Wrist-worn light loggerse can be easily covered by clothing or bedding [27-31]. At the same time, light loggers placed in the corneal plane (e.g., [32-36]) are not readily available for use in every-day contexts [37, 38].* | |

| | | |
|---|---|---|
| R1.5 | Isn't it necessary to discuss light exposure using circadian rhythm parameters? | Light exposure is of course a continuous 24-hour variable that needs to be considered over time. We have therefore added the qualifier “24 hours” where we sat fitting. |

| **Reviewer 2** | | | |
|---|---|---|---|
| **Comment** | **Reviewer comment** | **Response** | **Related comments** |
| R2.1 | As the field of light exposure research faces significant methodological and infrastructure gaps that limit progress and comparability across studies, establishing standardized tools, metrics, and protocols is essential to ensure reliable exposure assessment and facilitate the translation of results into health and public policy contexts. These efforts will strengthen evidence-based recommendations and promote the integration of light as a key determinant of human health. I believe that the effort to produce this consensus can provide guidance for future research and policy development, and for this reason, the article is valuable and interesting. However, some points deserve further attention, making it even more useful and suitable for multiple purposes. Here are some suggestions/observations: | Many thanks for your feedback and your thoughtful comments. | |
| R2.2 | Methods: The text states that there were 13 participants, but does not clearly specify the selection criteria, geographic distribution, or disciplinary diversity (how many were from public health, engineering, chronobiology, etc.). This may reduce the reproducibility and representativeness of the results. Given the limited population coverage of the representatives, it would be important to specify the selection criteria here and, in the study's limitations, point out the lack of representation of populations from different countries (such as low-income, non-European, other ancestries, and mixed race, among other factors). | We have now extended the section *Participants* in the *Methods* section:<br><br>*Thirteen experts (all co-authors of this paper) participated in the development process. Experts were purposively selected to ensure broad coverage of disciplinary domains relevant to non-visual effects of light, including chronobiology, vision science, public health, lighting engineering, radiation protection, and occupational health. Eligibility was based on demonstrated domain expertise (e.g., peer-reviewed publications, contributions to standards or guidelines, or applied professional experience). To facilitate in-depth discussion and iterative consensus building, the group size was intentionally limited to thirteen participants, spanning career stages from graduate student to full professor. All experts were* | |

| | | |
|---|---|---|
| | | *affiliated with European institutions; as such, while the group reflects disciplinary diversity, it does not represent a comprehensive geographic or demographic distribution.*<br><br>In the *Discussion*, we now also include a detailed *Limitations* section. |
| R2.3 | A description of the assessment of the consensus reached is also missing: there is no indication of whether the degree of agreement among experts was measured or how disagreements were resolved. | The consensus method is described in the section *Agreement and consensus*. |
| R2.4 | The article is extensive and technical, and non-specialist readers may have difficulty following it. Some parts could be simplified or presented in supplementary tables/figures. Perhaps an integrative conceptual figure showing the relationships between gaps, priorities, and capacity building could be added. | We thank the reviewer for this suggestion, and have added Figure 1 summarizing the findings of this workshop. |
| R2.5 | Perhaps this is the most relevant aspect of the article, which raises a question: the authors argue that positioning the article from a European perspective will fill important gaps and provide a solid foundation (page 17; Table 3 – "Positioning this work from a European perspective could therefore help address critical knowledge gaps and also provide a solid foundation…"). However, they also emphasize the idea that this is a global effort and need, while also presenting a geographic and institutional bias, given that most references and examples come from European countries. Since the text aims to establish a broad agenda, it would be helpful to acknowledge the lack of global representation and highlight the need to propose strategies that involve underrepresented regions (such as Asia, Latin America, and Africa). | We thank the reviewer for this comment. We have added the following text:<br><br>*At the same time, the field must move toward greater global inclusion by actively engaging underrepresented regions such as Asia, Latin America, and Africa, thereby ensuring that emerging frameworks and datasets capture diverse cultural and environmental realities.* |
| R2.6 | Although item 7 in the conclusions points to the lack of data on vulnerable populations, populations in other parts of the world are not necessarily vulnerable, but they are different from those in Europe. Highlighting this limitation and including the rest of the world in future | We have corrected Gap 7, which is now called: "Poor characterization of globally presentative and vulnerable populations" |

| | |
|---|---|
| | consensus will be of great value to this field. This concern should be included in the article. |

**New content and materials**
In addition to the reviewer comments, we have also made the following changes to our submission, which have strengthened the manuscript:

| Section | Description |
|---|---|
| Figures | We have added Figure 1, which shows the gaps and priorities succintly. |
